# Diagnostic Performance and Characteristics of Anterior Nasal Collection for the SARS-CoV-2 Antigen Test: A Prospective Study in Japan

**DOI:** 10.1101/2021.03.03.21252425

**Authors:** Yuto Takeuchi, Yusaku Akashi, Daisuke Kato, Miwa Kuwahara, Shino Muramatsu, Atsuo Ueda, Shigeyuki Notake, Koji Nakamura, Hiroichi Ishikawa, Hiromichi Suzuki

## Abstract

We conducted a prospective study in Japan to evaluate the diagnostic performance of the antigen test QuickNavi-COVID19 Ag using anterior nasal samples and to compare the degrees of coughs or sneezes induction and the severity of pain between anterior nasal collection and nasopharyngeal collection. A total of 862 participants were included in the analysis. In comparison to the findings of reverse transcription PCR using nasopharyngeal samples, the antigen test using anterior nasal samples showed 72.5% sensitivity (95% confidence interval [CI]: 58.3%-84.1%) and 100% specificity (95% CI: 99.3%-100%). Anterior nasal collection was associated with a significantly lower degree of coughs or sneezes induction and the severity of pain in comparison to nasopharyngeal collection (*p* < 0.001). The antigen test using anterior nasal samples showed moderate sensitivity but was less painful and induced fewer coughs or sneezes.

## INTRODUCTION

The laboratory diagnosis of severe acute respiratory syndrome coronavirus 2 (SARS-CoV-2) includes nucleic acid amplification tests (NAATs), antigen tests, and antibody tests (1). Antigen tests are generally less sensitive than NAATs for detecting SARS-CoV-2 but are relatively inexpensive, and most can be performed at the point of care (2).

The diagnostic performance of antigen test of nasopharyngeal samples has been evaluated, and the reported specificity is consistently high > 97%, while sensitivity ranges from 0% to 94% (2). We previously evaluated the performance of an antigen test, QuickNavi-COVID19 Ag (Denka Co., Ltd., Tokyo, Japan), using nasopharyngeal samples from 1186 participants (3). The overall sensitivity and specificity were 86.7% (95% confident interval [CI]: 78.6%-92.5%) and 100% (95% CI: 99.7%-100%), respectively, and the sensitivity for symptomatic participants was 91.7% (95% CI: 82.7%-96.9%) (3).

In recent studies, the sensitivity of anterior nasal samples was reported to be comparable to that of nasopharyngeal samples for NAATs (4, 5). Anterior nasal collection was reported to be less painful than nasopharyngeal collection (6) and may cause less droplet dispersal. These benefits are also expected in antigen tests but have yet to be fully evaluated.

We prospectively evaluated the diagnostic performance of the QuickNavi-COVID19 Ag test using anterior nasal samples and compared the degrees of coughs or sneezes induction and the severity of pain between anterior nasal collection and nasopharyngeal collection.

## MATERIALS AND METHODS

We conducted the present prospective study between October 7, 2020 and January 9, 2021, at a drive-through PCR center where participants were referred from a local public health center and 97 primary care facilities in Tsukuba, Japan. After receiving the participants’ informed consent, additional anterior nasal samples for the antigen test were collected and their clinical information was obtained. Cases with no clinical data were excluded from this study. In cases where participants enrolled in the current study more than once, only the first evaluation was included in this study. The present study was approved by the ethics committee of Tsukuba Medical Center Hospital (approval number: 2020-033).

### Sample collection and the antigen test procedure

We simultaneously obtained an anterior nasal sample for the antigen test and a nasopharyngeal sample for the PCR examination. All samples were obtained with FLOQSwabs (Copan Italia S.p.A., Brescia, Italy).

The anterior nasal sample was initially collected according to the manufacturer’s instructions. Namely, a nasopharyngeal-type flocked (NP-type) swab was inserted to 2 cm depth in one nasal cavity, rotated five times, and held in place for five seconds. The antigen test using the QuickNavi-COVID19 Ag kit was performed immediately after anterior nasal collection and the result was obtained by the visual interpretation of each examiner.

A nasopharyngeal sample was subsequently collected with an NP-type swab according to a previously described procedure (7) and was diluted in 3 mL of Universal Transport Medium (UTM) (Copan Italia S.p.A., Brescia, Italy). The UTM was transferred to an in-house microbiology laboratory located next to the drive-through sample-collecting site of the PCR center within an hour of sample collection.

### SARS-CoV-2 detection using reverse transcription PCR

After the arrival of the UTM samples, purification and RNA extraction were performed with magLEAD 6gC (Precision System Science Co., Ltd., Chiba, Japan) from 200 µL aliquots of UTM for in-house reverse transcription (RT)-PCR on the same day as sample collection. RNA was eluted in 100 µL and stored at-80 °C after the in-house RT-PCR test. The eluted samples were transferred to Denka Co., Ltd., every week for a reference real-time RT-PCR test on Applied Biosystems QuantStudio 3 (Thermo Fisher Scientific Inc., Waltham, MA, USA) using a QuantiTect Probe RT-PCR Kit (QIAGEN Inc., Germantown, MD, USA) and primer/probe N and N2 set (8).

The presence or absence of SARS-CoV-2 was defined by the results of the reference real-time RT-PCR test. However, if discordance existed between the reference real-time RT-PCR test and the in-house RT-PCR test, a re-evaluation was performed with an Xpert Xpress SARS-CoV-2 and GeneXpert System (Cepheid, Sunnyvale, CA, USA), the results of which provided the final judgment.

### The degrees of coughs or sneezes and the severity of pain induced by the sample collection procedure

The degrees of coughs or sneezes and the severity of pain caused by the insertion of the swab into the anterior nasal cavity and nasopharynx in the same participant were assessed. Examiners rated the degrees of coughs or sneezes induction from the following four categories: “None”, “Small, 1-2 times”, “Loud, 1-2 times” and “Loud, multiple times”. The severity of pain was evaluated with a five-point scale (Pain score), with 1 being “no pain” and 5 being“worst imaginable pain,” and the participants were asked to report a number from the scale.

### The comparison of SARS-CoV-2 viral loads between different sample collection sites and swab types

We conducted an additional experiment to evaluate whether the viral loads differed between sample collection sites and swab types between January 8 and 19, 2021. After receiving the participants’ informed consent, two anterior nasal samples were obtained from the participants for whom a nasopharyngeal sample had already tested positive for SARS - CoV-2. Two anterior nasal swab samples were collected from each nostril using one with a NP-type swab and the other with an oropharyngeal-type flocked (OP-type) swab. These sample collections were performed on the same day.

The samples were diluted in 3 mL of UTM, and stored at −80°C. After several days of storage, the samples were thawed, and purification and RNA extraction were performed according to the above-described method. The viral concentrations in samples were quantified with the following procedure. The calibration curves were generated with 5, 50, and 500 copies/reaction of positive control (EDX SARS-CoV-2 Standard; Bio-Rad Laboratories, Inc., Hercules, CA, USA). Quantitative RT-PCR was performed on a LightCycler 96 System (Roche, Basel, Switzerland) using a THUNDERBIRD Probe One-step qRT-PCR Kit (TOYOBO Co. Ltd., Osaka, Japan) with a primer/probe N2 set.

### Statistical analyses

The sensitivity, specificity, positive predictive value (PPV), and negative predictive value (NPV) of QuickNavi-COVID19 Ag and their 95% confidence intervals (CIs) were calculated with the Clopper and Pearson method. The viral loads according to collection sites and swab types were compared by the Wilcoxon signed-rank test with Holm correction. The degrees of coughs or sneezes induction and the pain score were also compared between the two different collection procedures using the the McNemar-Bowker test and the Wilcoxon signed-rank test, respectively. *P* values of < 0.05 were considered to indicate statistical significance. All statistical analyses were conducted using the R 3.5.2 software program (The R Foundation, Vienna, Austria).

## RESULTS

### Viral loads in different sample collection sites and swab types

In 32 identical SARS-CoV-2 positive cases, we evaluated the SARS-CoV-2 viral loads of nasopharyngeal samples (NPS), anterior nasal samples with NP-type swabs (AWN), and anterior nasal samples with OP-type swabs (AWO) (Fig. 1). The median viral loads for NPS, AWN, and AWO were 53,560 (interquartile range [IQR]: 605-608,050), 1,792 (IQR: 7-81,513), and 6,369 (IQR: 7-97,535), respectively. With the NPS as a reference, the PCR-positive rate for AWN was 84.4% (27/32;95% CI: 67.2%-94.7%), while that for AWO was 81.3% (26/32; 95% CI: 63.6%-92.8%). In comparison to NPS, the viral load was significantly lower with AWN (*p* < 0.001) and AWO (*p* < 0.001), but there was no significant difference between AWN and AWO (*p* = 0.61). The viral loads and Ct values for all samples are described in Table S1a and Table S1b.

**FIG 1.**
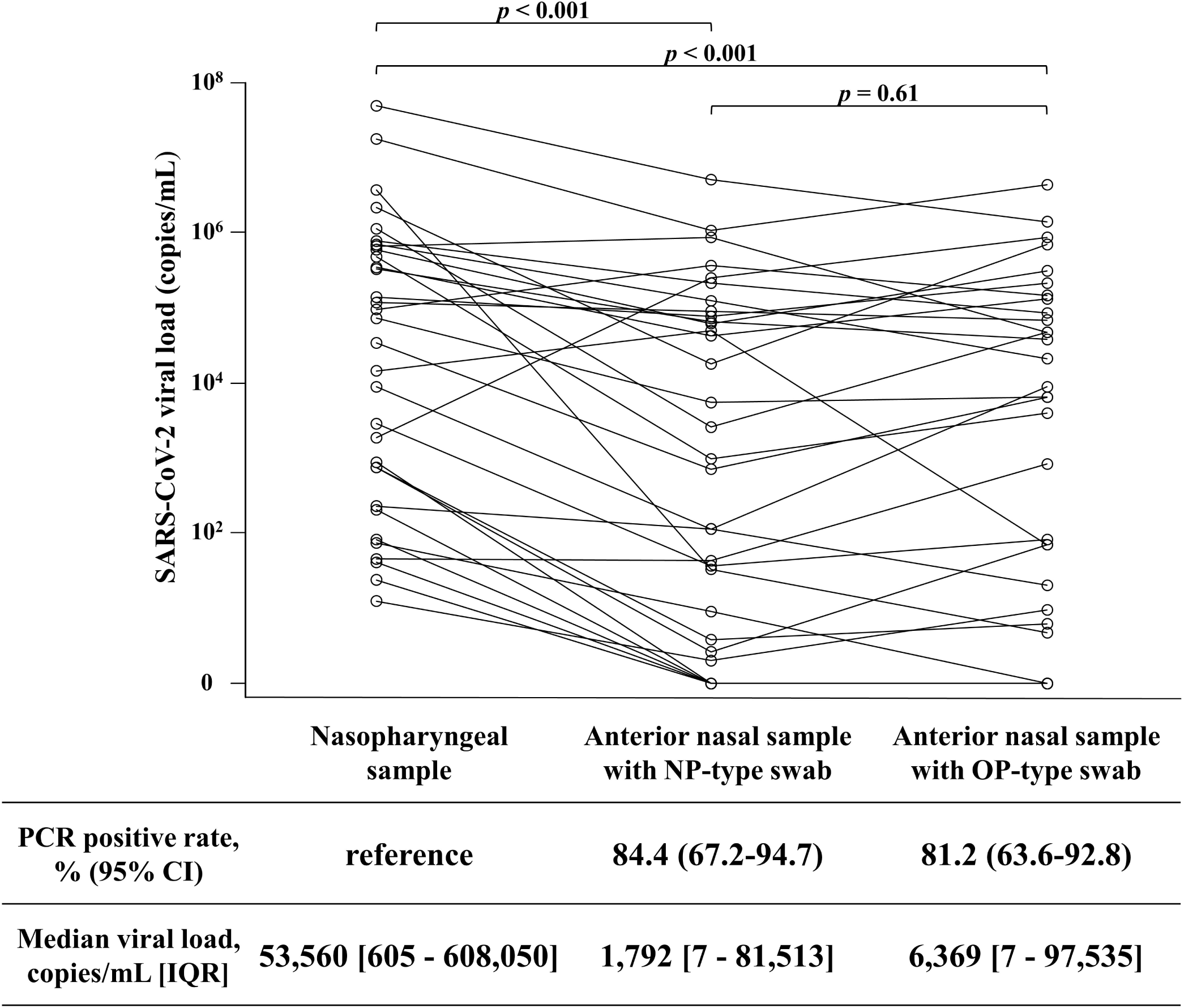
Comparison of SARS-CoV-2 viral loads between each collection site and swab type. The set of viral loads in the same participant is represented by connected lines. The viral loads were compared by the Wilcoxon signed-rank test, and *P* values are corrected with Holm correction. NP swab, nasopharyngeal-type swab; OP swab, oropharyngeal-type swab; CI, confidence interval; IQR, interquartile range.

### Demographic data of study population

A total of 876 participants were screened for the evaluation. Most samples were obtained at the drive-through PCR center, and only 17 were obtained after hospitalization. We excluded the participants who underwent duplicate tests (n=7) or for whose clinical information were lacking (n=7). Finally, 862 participants were included in the analysis.

Among the 862 participants, SARS-CoV-2 was detected in 50 (5.8%) on nasopharyngeal samples by the reference real-time RT-PCR test. There was one discordant sample that was positive on the in-house RT-PCR test and negative on the reference real-time RT-PCR test. The Xpert Xpress SARS-CoV-2 test provided a positive result for the sample (cycle threshold [Ct] values for E and N2 targets: 0.0 and 39.8, respectively); thus, 51 (5.9%) participants were finally considered positive for SARS-CoV-2. The discordant sample was obtained from a participant who had been diagnosed with COVID-19 one month before the current evaluation and who was referred to the PCR center due to refractory respiratory symptoms.

All 51 participants who were positive for SARS-CoV-2 were symptomatic (Table 1a); their characteristics are described in Table 1b. The most common symptom was fever (80.4%), followed by cough or sputum production (60.8%), sore throat (37.3%), runny nose or nasal congestion (35.5%), and loss of taste or smell (27.5%).

**Table 1a.** Demographic data of the whole study population and cases infected with SARS-CoV-2

|  | Total | SARS-CoV-2 |  |
| --- | --- | --- | --- |
|  |  | Positive | Negative |
| N | 862 | 51 | 811 |
| Age (years, median [IQR]) | 36.0 [24.0, 48.0] | 43.0 [30.0, 57.5] | 35.0 [23.0, 47.0] |
| <18 years | 106 (12.3) | 2 (3.9) | 104 (12.8) |
| 18-64 years | 678 (78.7) | 44 (86.3) | 634 (78.2) |
| ≥ 65 years | 78 (9.0) | 5 (9.8) | 73 (9.0) |
| Sex (Female) | 383 (44.4) | 19 (37.3) | 364 (44.9) |
| Asymptomatic participants | 72 (8.4) | 0 (0.0) | 72 (8.9) |
| Symptomatic participants | 790 (91.6) | 51 (100) | 739 (91.1) |

**Table 1b.** Characteristics of symptomatic participants and cases infected with SARS-CoV-2

|  | Total | SARS-CoV-2 |  |
| --- | --- | --- | --- |
|  |  | Positive | Negative |
| N | 790 | 51 | 739 |
| Days from symptom onset to sample collection [IQR] | 2.0 [1.0, 3.0] | 3.0 [2.0, 5.0] | 2.0 [1.0, 3.0] |
| Signs and symptoms |  |  |  |
| Fever | 628 (79.5) | 41 (80.4) | 587 (79.4) |
| Cough/sputum production | 255 (32.3) | 31 (60.8) | 224 (30.3) |
| Runny nose/nasal congestion | 185 (23.4) | 18 (35.3) | 167 (22.6) |
| Loss of taste or smell | 32 (4.1) | 14 (27.5) | 18 (2.4) |
| Dyspnea | 7 (0.9) | 2 (3.9) | 5 (0.7) |
| Fatigue | 105 (13.3) | 10 (19.6) | 95 (12.9) |
| Diarrhea | 48 (6.1) | 1 (2.0) | 47 (6.4) |
| Sore throat | 210 (26.6) | 19 (37.3) | 191 (25.8) |
| Headache | 121 (15.3) | 11 (21.6) | 110 (14.9) |

### Diagnostic performance of QuickNavi-COVID19 Ag using anterior nasal samples

Of the 51 participants who were found to be SARS-CoV-2-positive by the RT-PCR test, 37 participants were found to be positive with the antigen test with anterior nasal samples (Table 2). Among the 811 SARS-CoV-2-negative participants, all participants were found to be negative with the antigen test (Table 2). The concordance rate between the antigen test and RT-PCR was thus 98.4% (95% CI: 97.3%-99.1%). The sensitivity, specificity, PPV, and NPV were 72.5% (95% CI: 58.3%-84.1%), 100% (95% CI: 99.3%-100%), 100% (95%CI: 86.2%-100%), and 98.3% (95%CI: 97.2%-99.1%), respectively (Table 2).

**Table 2.**
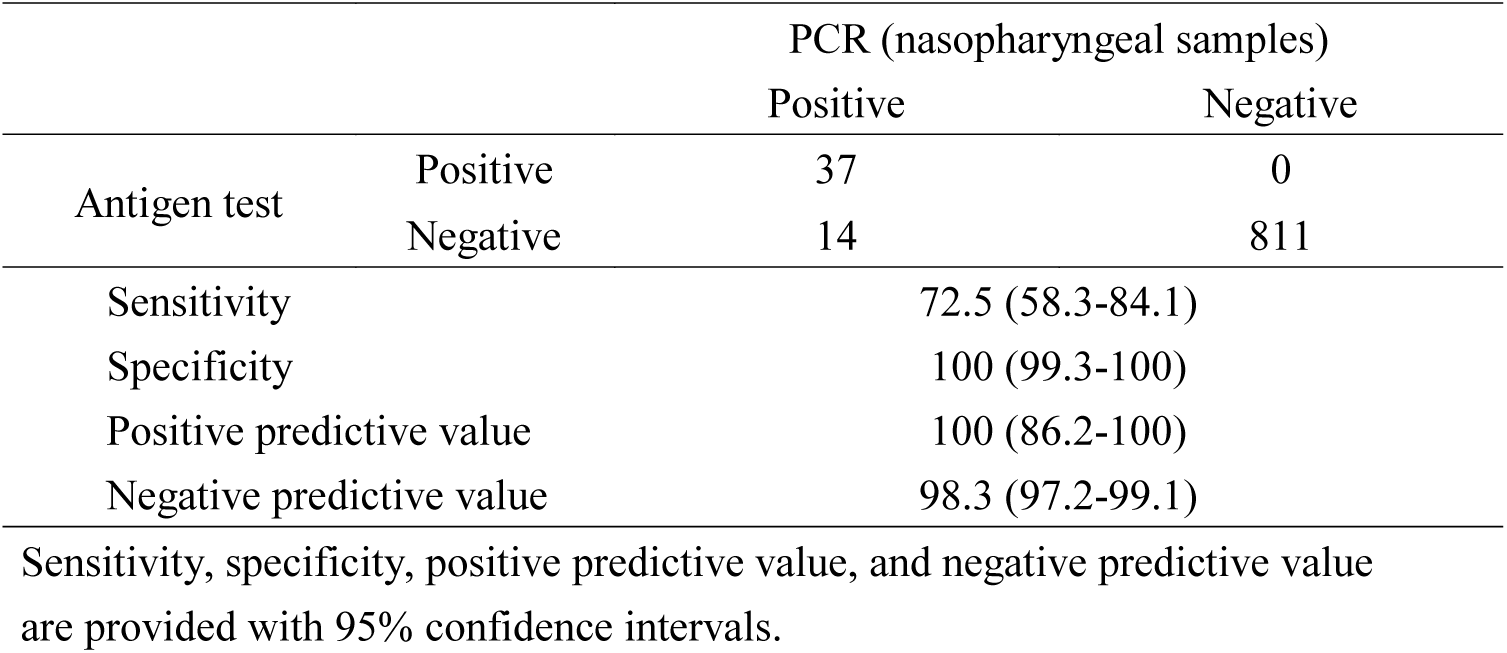
Clinical performance of antigen test using anterior nasal samples

|  |  | PCR (nasopharyngeal samples) |  |
| --- | --- | --- | --- |
|  |  | Positive | Negative |
| Antigen test | Positive | 37 | 0 |
|  | Negative | 14 | 811 |
| Sensitivity |  | 72.5 (58.3-84.1) |  |
| Specificity |  | 100 (99.3-100) |  |
| Positive predictive value |  | 100 (86.2-100) |  |
| Negative predictive value |  | 98.3 (97.2-99.1) |  |

Sensitivity, specificity, positive predictive value, and negative predictive value are provided with 95% confidence intervals.

### Comparison of degrees of coughs or sneezes induction and the severity of pain between anterior nasal and nasopharyngeal sample collection

The degrees of coughs or sneezes induced by sample collection was measured in 784 participants (Fig. 2). Coughing or sneezing was observed in 149 (19.0%) of anterior nasal collections and in 316 (40.3%) of nasopharyngeal collection. When coughs or sneezes occurred in anterior nasal collection, their degrees were significantly lower than in nasopharyngeal collection (*p* < 0.001).

**FIG 2.**
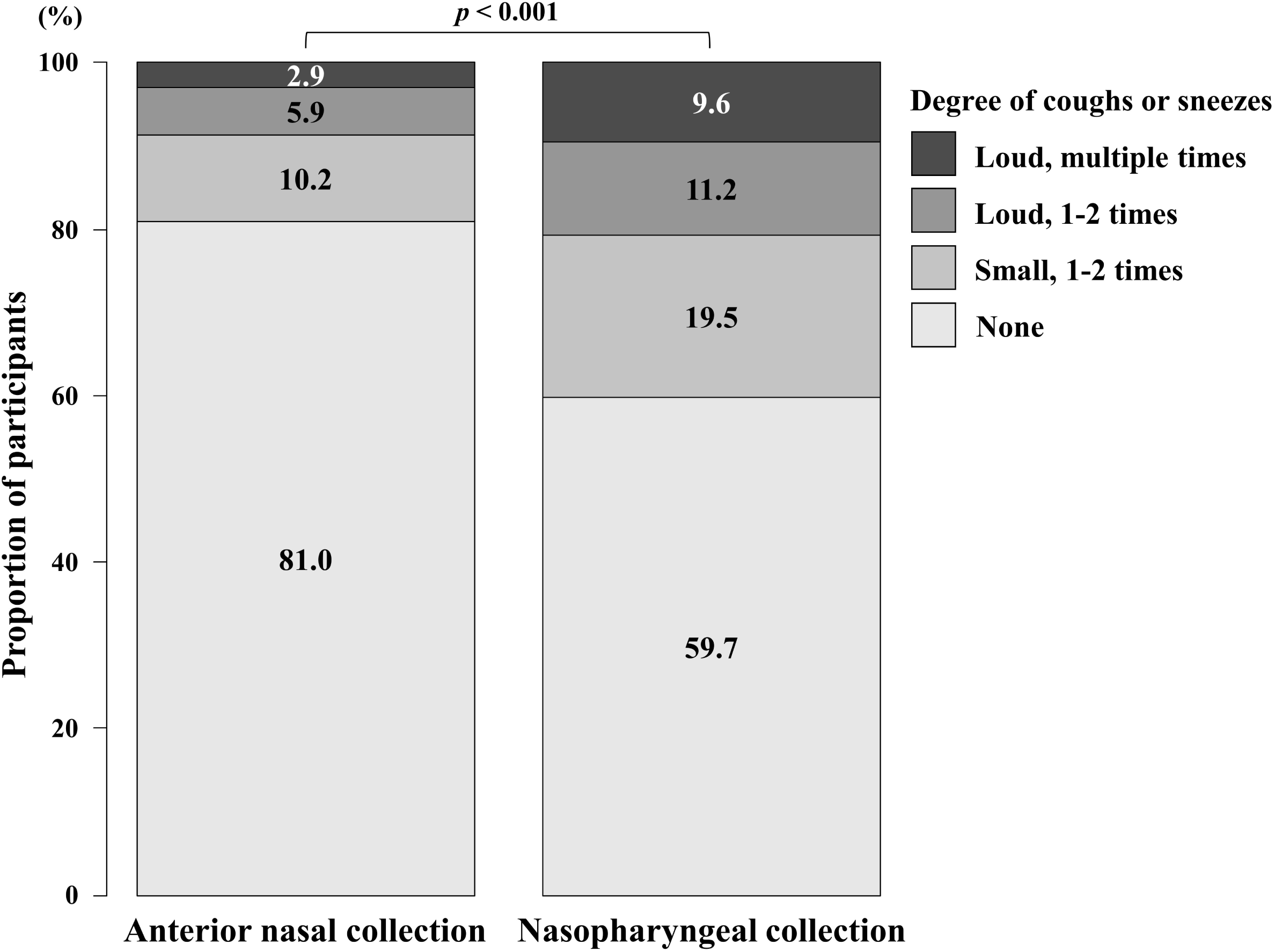
Degrees of coughs or sneezes induced by anterior nasal and nasopharyngeal collection. The degrees of coughing or sneezing following swab insertion was compared between anterior nasal collection and nasopharyngeal collection in the same participant (n = 784). The degrees were rated on a four-point scale. The McNemar-Bowker test was used for the comparison.

The pain score was obtained from 90 participants (Fig. 3). Fifty-seven participants (63.3%) reported no pain in anterior nasal collection. The median pain score of anterior nasal collection and nasopharyngeal collection was 1 (IQR: 1-2) and 3 (IQR: 2-4), respectively. In comparison to nasopharyngeal collection, anterior nasal collection was significantly less painful (*p* < 0.001).

**FIG 3.**
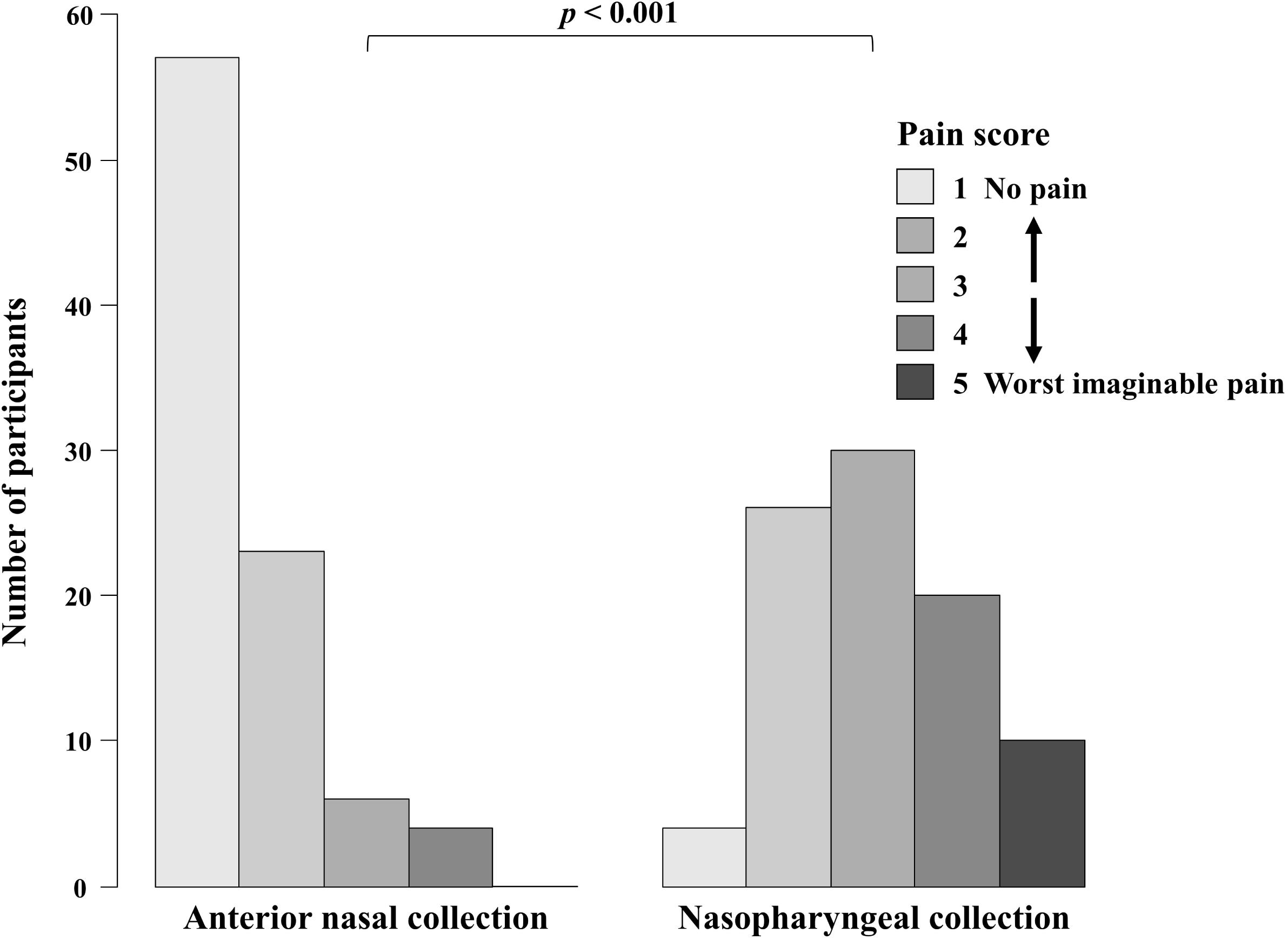
The pain score in anterior nasal collection and nasopharyngeal collection. The severity of pain at swab insertion was assessed on a five-point scale, from 1 to 5 (Pain score). The pain score for each collection method was obtained from the same participant (n = 90). The comparison of the pain scores with the two collection procedures was performed with the Wilcoxon signed-rank test.

## DISCUSSION

The QuickNavi-COVID19 Ag test using anterior nasal samples showed 72.5% sensitivity and 100% specificity. In comparison to nasopharyngeal collection, anterior nasal collection was less painful and was associated with fewer coughs or sneezes. In addition, the study demonstrated that the viral load of anterior nasal samples was significantly lower than that of nasopharyngeal samples. Meanwhile, the swab types did not influence the viral load of anterior nasal samples.

In the current study, anterior nasal samples provided a lower antigen test sensitivity than our previous study evaluating nasopharyngeal samples (3). The sensitivity of antigen tests is largely influenced by the viral load in collected samples (9–12). The QuickNavi-COVID19 Ag test could detect SARS-CoV-2 in almost all samples with Ct values < 30, and in 18.8% of samples with Ct values > 30 (3). The viral load may vary between collection sites (13), and this study recognized the viral load of samples was significantly lower when they were collected from the anterior nasal cavity (Fig. 1). This lower viral load in anterior nasal samples could explain the decreased sensitivity. On the other hand, the QuickNavi-COVID19 Ag test provided 100% specificity in both the present study and our previous study (3). Although it is necessary to verify whether similar results can be obtained in other settings, false positives should be avoided to prevent unnecessary additional testing and inappropriate isolation measures (14).

We observed that anterior nasal collection caused fewer coughs or sneezes in comparison to nasopharyngeal sample collection (Fig. 2). Notably, anterior nasal collection induced coughs or sneezes in only 19% of participants. Coughs or sneezes generate droplets and prolong their dispersal by forming multiphase turbulent gas clouds (15), which leads to greater droplet exposure. SARS-CoV-2 is mainly transmitted through droplets (16); thus, anterior nasal collection, which was associated with fewer coughs or sneezes induction, may reduce the transmission risk among healthcare providers.

Anterior nasal collection was less painful (Fig. 3), with more than half of the participants reporting no pain from the procedure. Nasopharyngeal collection is an uncomfortable and painful experience (6) and may discourage patients from receiving tests. Besides, nasopharyngeal collection may not be applicable if patients have a history of recent nasal trauma or surgery, remarkable nasal septum deviation, or marked coagulopathy (7). Despite the decreased sensitivity, when NAATs are not readily available, an antigen test with anterior nasal samples may be an option in these clinical contexts.

The selection of swab type influences the uptake, extraction and recovery efficiency of the collected sample (17, 18). In this study, we compared two flocked swabs with different tip sizes (NP-type and OP-type swab). There was no significant difference in the viral load of the samples collected with the two types of swabs; however, the OP-type swab has a larger tip and seemed to handle a larger amount of samples collected. A previous study suggested that the efficiency of sample release was not associated with the absorbed volume (19), which could explain the result in this study.

The present study was associated with some limitations. First, the samples used for the reference real-time RT-PCR test were frozen and transported. Although the samples were frozen at −80°C, the viral loads may have been decreased during the storage and transport process. Nevertheless, in the case of discrepancy with in-house PCR, re-evaluation was performed and did not affect the calculation of the sensitivity of the antigen test. Second, asymptomatic SARS-CoV-2 positive patients were unintentionally not included in this study. Further study is required to evaluate the clinical performance of the antigen test in those patients. Finally, this study was conducted at a single center, although participants were referred from multiple primary facilities. Further research should be conducted to assess the generalizability of the findings.

In conclusion, the QuickNavi-COVID19 Ag test with an anterior nasal sample showed 100% specificity; however, the sensitivity in the detection of SARS-CoV-2 was lower in comparison to the QuickNavi-COVID19 Ag test with nasopharyngeal samples. Anterior nasal collection was less invasive and induced fewer coughs or sneezes, which may be more comfortable for the patient and may reduce the risk of droplet exposure to healthcare workers.

## Data Availability

The data are not publicly available due to their containing information that could compromise the privacy of research participants.

## ACKNOWLEDGEMENTS

We thank Mrs. Yoko Ueda, Mrs. Mio Matsumoto, Dr. Yumi Hirose and the staff in the Department of Clinical Laboratory of Tsukuba Medical Center Hospital for their intensive support of this study. We thank all of the medical institutions for providing their patients’ clinical information. Mrs. Yoko Ueda and Mrs. Mio Matsumoto significantly contributed to creating the database of this study.

Denka Co., Ltd., provided fees for research expenses and the QuickNavi-COVID19 Ag kits without charge. Hiromichi Suzuki received a lecture fee from Otsuka Pharmaceutical Co., Ltd., regarding this study. Daisuke Kato, Miwa Kuwahara and Shino Muramatsu belong to Denka Co., Ltd., the developer of the QuickNavi-COVID19 Ag.

## Notes

### Author Declarations

The ethics committee of TMCH approved the present study (approval number: 2020-033).

## REFERENCES

1. World Health Organization. 2020. Diagnostic testing for SARS-CoV-2: interim guidance, 11 September 2020. https://apps.who.int/iris/handle/10665/334254. Accessed 24 February 2021.

2. World Health Organization. 2020. Antigen -detection in the diagnosis of SARS-CoV-2 infection using rapid immunoassays: interim guidance, 11 September 2020. https://apps.who.int/iris/handle/10665/334253. Accessed 24 February 2021.

3. Takeuchi Y, Akashi Y, Kato D, Kuwahara M, Muramatsu S, Ueda A, Notake S, Nakamura K, Ishikawa H, Suzuki H. 2021. The evaluation of a newly developed antigen test (QuickNavi ™ COVID19 Ag) for SARS-CoV-2: A prospective observational study in Japan. medRxiv https://doi.org/10.1101/2020.12.27.20248876.

4. Péré H, Podglajen I, Wack M, Flamarion E, Mirault T, Goudot G, Hauw-Berlemont C, Le L, Caudron E, Carrabin S, Rodary J, Ribeyre T, Bélec L, Veyer D. 2020. Nasal Swab Sampling for SARS-CoV-2: a Convenient Alternative in Times of Nasopharyngeal Swab Shortage. J Clin Microbiol https://doi.org/10.1128/jcm.00721 - 20.

5. Tu Y-P, Jennings R, Hart B, Cangelosi GA, Wood RC, Wehber K, Verma P, Vojta D, Berke EM. 2020. Swabs Collected by Patients or Health Care Workers for SARS - CoV-2 Testing. N Engl J Med 383:494 –496.

6. Frazee BW, Rodríguez-Hoces de la Guardia A, Alter H, Chen CG, Fuentes EL, Holzer AK, Lolas M, Mitra D, Vohra J, Dekker CL. 2018. Accuracy and Discomfort of Different Types of Intranasal Specimen Collection Methods for Molecular Influenza Testing in Emergency Department Patients. Ann Emerg Med 71:509 - 517.e1.

7. Marty FM, Chen K, Verrill KA. 2020. How to Obtain a Nasopharyngeal Swab Specimen. N Engl J Med 382:e76.

8. Shirato K, Nao N, Katano H, Takayama I, Saito S, Kato F, Katoh H, Sakata M, Nakatsu Y, Mori Y, Kageyama T, Matsuyama S, Takeda M. 2020. Develop ment of genetic diagnostic methods for detection for novel coronavirus 2019(nCoV-2019) in Japan. Jpn J Infect Dis 73:304 –307.

9. Hirotsu Y, Maejima M, Shibusawa M, Amemiya K, Nagakubo Y, Hosaka K, Sueki H, Hayakawa M, Mochizuki H, Tsutsui T, Kakizaki Y, Miyashita Y, Omata M. 2021. Prospective Study of 1,308 Nasopharyngeal Swabs from 1,033 Patients using the LUMIPULSE SARS-CoV-2 Antigen Test: Comparison with RT -qPCR. Int J Infect Dis https://doi.org/10.1016/j.ijid.2021.02.005.

10. Aoki K, Nagasawa T, Ishii Y, Yagi S, Okuma S, Kashiwagi K, Maeda T, Miyazaki T, Yoshizawa S, Tateda K. 2021. Clinical validation of quantitative SARS-CoV-2 antigen assays to estimate SARS-CoV-2 viral loads in nasopharyngeal swabs. J Infect Chemother https://doi.org/10.1016/j.jiac.2020.11.021.

11. Salvagno GL, Gianfilippi G, Bragantini D, Henry BM, Lippi G. 2021. Clinical assessment of the Roche SARS-CoV-2 rapid antigen test. Diagnosis https://doi.org/10.1515/dx-2020-0154.

12. Merino P, Guinea J, Muñoz-Gallego I, González-Donapetry P, Galán JC, Antona N, Cilla G, Hernáez-Crespo S, Díaz-de Tuesta Jl, Torrella AG, González-Romo F, Escribano P, Sánchez-Castellano MÁ, Sota-Busselo M, Delgado-Iribarren A, García J, Cantón R, Muñoz P, Folgueira MD, Cuenca-Estrella M, Oteo-Iglesias J, Medrano S, Pérez A, Galar A, Martínez-Expósito O, Alejo-Cancho I, Martín-Higuera Mc, Rolo M, Estévez MJ, Bravo T, Vicente D, Montes M. 2021. Multicenter evaluation of the Panbio™ COVID-19 rapid antigen-detection test for the diagnosis of SARS - CoV-2 infection. Clin Microbiol Infect https://doi.org/10.1016/j.cmi.2021.02.001.

13. Lee RA, Herigon JC, Benedetti A, Pollock NR, Denkinger CM. 2021. Performanceof Saliva, Oropharyngeal Swabs, and Nasal Swabs for SARS-CoV-2 Molecular Detection: A Systematic Review and Meta-analysis. J Clin Microbiol https://doi.org/10.1128/jcm.02881-20.

14. Ogawa T, Fukumori T, Nishihara Y, Sekine T, Okuda N, Nishimura T, Fujikura H, Hirai N, Imakita N, Kasahara K. 2020. Another false -positive problem for a SARS - CoV-2 antigen test in Japan. J Clin Virol 131:104612.

15. Bourouiba L. 2020. Turbulent Gas Clouds and Respiratory Pathogen Emissions: Potential Implications for Reducing Transmission of COVID −19. JAMA - J Am Med Assoc 323:1837–1838.

16. World Health Organization. 2020. Transmission of SARS-CoV-2 : implications for infection prevention precautions. https://www.who.int/publications/i/item/modes-of-transmission-of-virus-causing-covid-19-implications-for-ipc-precaution-recommendations. Accessed 24 February 2021.

17. Kahamba TR, Noble L, Stevens W, Scott L. 2020. Comparison of three nasopharyngeal swab types and the impact of physiochemical properties for optimal SARS-CoV-2 detection. medRxiv https://doi.org/10.1101/2020.10.21.20206078.

18. Bruijns BB, Tiggelaar RM, Gardeniers H. 2018. The Extraction and Recovery Efficiency of Pure DNA for Different Types of Swabs. J Forensic Sci 63:1492 –1499.

19. Zasada AA, Zacharczuk K, Woźnica K, Główka M, Ziółkowski R, Malinowska E. 2020. The influence of a swab type on the results of point -of-care tests. AMB Express https://doi.org/10.1186/s13568-020-00978-9.

