## Supplementary material for "Diagnostic Performance and Characteristics of Anterior Nasal Collection for the SARS-CoV-2 Antigen Test: A Prospective Study in Japan": Table S1a, Table S1b

Table S1a. SARS-CoV-2 viral load for each sampling site and swab type

| Case number | Viral load (copies/mL) | | |
|---|---|---|---|
| | Nasopharyngeal sample | Anterior nasal sample with NP-type swab | Anterior nasal sample with OP-type swab |
| 1 | 755 | 3 | 5 |
| 2 | 94930 | 357700 | 145400 |
| 3 | 1804 | 255800 | 851300 |
| 4 | 730 | 2 | 70 |
| 5 | 490700 | 962 | 4018 |
| 6 | 45 | 42 | 818 |
| 7 | 2851 | 35 | 82 |
| 8 | 340000 | 41570 | 133100 |
| 9 | 73090 | 5532 | 6415 |
| 10 | 14180 | 49480 | 70 |
| 11 | 2139000 | 18120 | 683000 |
| 12 | 706800 | 126100 | 21140 |
| 13 | 588900 | 63240 | 310800 |
| 14 | 136700 | 78300 | 216900 |
| 15 | 34030 | 688 | 6322 |
| 16 | 8616 | 114 | 8763 |
| 17 | 207 | Not detected | Not detected |
| 18 | 119400 | 91150 | 69960 |
| 19 | 228 | 109 | 19 |
| 20 | 791900 | 213300 | 85680 |
| 21 | 886 | Not detected | Not detected |
| 22 | 71 | 8 | Not detected |
| 23 | 3773000 | 31 | 4 |
| 24 | 665500 | 855800 | 48210 |
| 25 | 327400 | 66120 | 37970 |
| 26 | 22 | Not detected | Not detected |
| 27 | 40 | Not detected | Not detected |
| 28 | 79 | Not detected | Not detected |
| 29 | 48670000 | 4994000 | 1386000 |
| 30 | 11 | 1 | 8 |
| 31 | 17480000 | 1047000 | 4341000 |
| 32 | 1119000 | 2621 | 48130 |

NP-type, nasopharyngeal-type; OP-type, oropharyngeal-type.

Table S2b. SARS-CoV-2 cycle threshold (Ct) value for each sampling site and swab type

| Case number | Ct value | | |
|---|---|---|---|
| | Nasopharyngeal sample | Anterior nasal sample with NP-type swab | Anterior nasal sample with OP-type swab |
| 1 | 27.9 | 35.8 | 34.8 |
| 2 | 16.6 | 17.9 | 19.2 |
| 3 | 24.8 | 18.4 | 16.5 |
| 4 | 27.3 | 35.9 | 30.8 |
| 5 | 18.3 | 27.2 | 25.3 |
| 6 | 31.6 | 31.5 | 27.4 |
| 7 | 25.6 | 31.9 | 30.7 |
| 8 | 18.9 | 21.8 | 20.2 |
| 9 | 21.0 | 24.7 | 24.5 |
| 10 | 21.9 | 19.7 | 31.2 |
| 11 | 13.1 | 21.5 | 15.1 |
| 12 | 15.1 | 18.1 | 21.2 |
| 13 | 15.4 | 19.3 | 16.5 |
| 14 | 18.0 | 19.0 | 17.1 |
| 15 | 20.4 | 27.4 | 23.4 |
| 16 | 22.9 | 30.6 | 22.8 |
| 17 | 29.5 | Not detected | Not detected |
| 18 | 18.2 | 18.7 | 19.2 |
| 19 | 29.3 | 30.6 | 33.7 |
| 20 | 14.8 | 17.2 | 18.8 |
| 21 | 26.9 | Not detected | Not detected |
| 22 | 34.4 | 34.3 | Not detected |
| 23 | 14.0 | 32.1 | 35.5 |
| 24 | 16.7 | 16.3 | 20.8 |
| 25 | 17.8 | 20.3 | 21.1 |
| 26 | 32.3 | Not detected | Not detected |
| 27 | 31.5 | Not detected | Not detected |
| 28 | 30.6 | Not detected | Not detected |
| 29 | 12.5 | 15.6 | 17.3 |
| 30 | 33.2 | 36.5 | 33.6 |
| 31 | 13.9 | 17.7 | 15.8 |
| 32 | 17.6 | 25.9 | 21.9 |

NP-type, nasopharyngeal-type; OP-type, oropharyngeal-type.
